# Hydroxychloroquine alone or in combination with azithromycin to prevent major clinical events in hospitalised patients with coronavirus infection (COVID-19): rationale and design of a randomised, controlled clinical trial

**DOI:** 10.1101/2020.05.19.20106997

**Authors:** Alexandre B Cavalcanti, Fernando G Zampieri, Luciano CP Azevedo, Regis Rosa, Álvaro Avezum, Viviane C Veiga, Renato D. Lopes, Letícia Kawano-Dourado, Lucas P Damiani, Adriano J Pereira, Ary Serpa-Neto, Remo Furtado, Bruno Tomazini, Fernando A Bozza, Israel S. Maia, Maicon Falavigna, Thiago C Lisboa, Henrique Fonseca, Flávia R Machado, Otavio Berwanger, for the COALITION COVID-19 Brasil I Investigators

## Abstract

**Introduction:** Hydroxychloroquine and its combination with azithromycin have been suggested to improve viral clearance in patients with COVID-19, but its effect on clinical outcomes remains uncertain.

**Methods and analysis:** We describe the rationale and design of an open-label pragmatic multicentre randomised (concealed) clinical trial of 7 days of hydroxychloroquine (400 mg BID) plus azithromycin (500 mg once daily), hydroxychloroquine 400 mg BID, or standard of care for moderately severe hospitalised patients with suspected or confirmed COVID-19 (in-patients with up to 4L/minute oxygen supply through nasal catheter). Patients are randomised in around 50 recruiting sites and we plan to enrol 630 patients with COVID-19. The primary endpoint is a 7-level ordinal scale measured at 15-days: 1)not hospitalised, without limitations on activities; 2)not hospitalised, with limitations on activities; 3)hospitalised, not using supplementary oxygen; 4)hospitalised, using supplementary oxygen; 5)hospitalised, using high-flow nasal cannula or non-invasive ventilation; 6)hospitalised, on mechanical ventilation; 7)death. Secondary endpoints are the ordinal scale at 7 days, need for mechanical ventilation and rescue therapies during 15 days, need of high-flow nasal cannula or non-invasive ventilation during 15 days, length of hospital stay, in-hospital mortality, thromboembolic events, occurrence of acute kidney injury, and number of days free of respiratory support at 15 days. Secondary safety outcomes include prolongation of QT interval on electrocardiogram, ventricular arrhythmias, and liver toxicity. The main analysis will consider all patients with confirmed COVID-19 in the groups they were randomly assigned.

**Ethics and dissemination:** This study has been approved by Brazil’s National Ethic Committee (CONEP) and National Health Surveillance Agency (ANVISA). An independent data monitoring committee will perform interim analyses and evaluate adverse events throughout the trial. Results will be submitted for publication after enrolment and follow-up are complete, as well as presented and reported to local health agencies.

**ClinicalTrials.gov identifier:** NCT04322123

Strengths and limitations of this study
- Pragmatic randomised controlled trial of 7 days of hydroxychloroquine plus azithromycin, hydroxychloroquine or standard of care for moderately severe in-patients with suspected or confirmed COVID-19
- Multicentre: around 50 recruiting sites in Brazil with planned enrolment of 630 patients (1:1:1)
- The primary endpoint is a 7-level ordinal scale ([1] not hospitalised, without limitations on activities; [2] not hospitalised, with limitations on activities; [3] hospitalised, not using supplementary oxygen; [4] hospitalised, using supplementary oxygen; [5] hospitalised, using high-flow nasal cannula or non-invasive ventilation; [6] hospitalised, on mechanical ventilation; [7] death) measured at 15 days.
- Open label design (no placebo)

## Introduction

The novel coronavirus (SARS-CoV-2) disease (COVID-19) lacks targeted therapies. New therapies that minimize symptoms, reduce contagion time and, mainly, reduce complications and mortality are of public interest and should be studied in controlled settings, within scientific rigor.

Preliminary and low level of evidence suggests that hydroxychloroquine (HCQ) might be active against COVID-19 [1-4]. However, observational studies and small randomised trials have not found any benefit [5-8]. Furthermore, chloroquine and its derivates are associated with several possible side effects, such as hypoglycaemia, QT prolongation and cardiac toxicity [9]. Therefore, its use for this indication outside research protocols should be discouraged [10]. Likewise, the use of azithromycin appears as a potential adjuvant drug to hydroxychloroquine, with its open-label and non-randomised use associated with lower viral loads in patients with COVID-19 [2]. Nevertheless, its potential clinical benefits remain uncertain.

We are conducting a pragmatic randomised, open-label clinical trial, comparing standard treatment versus standard treatment added to 7 days of hydroxychloroquine plus azithromycin (800 mg and 500 mg daily, respectively) or hydroxychloroquine (800 mg daily) to prevent respiratory complications in patients admitted with suspected COVID19 pneumonia. We used the Standard Protocol Items: Recommendations for Interventional Trials (SPIRIT) guideline for this report, which is presented in the Electronic Supplementary File (ESM) [11]. The steering committee members are shown in the electronic supplementary material (ESM) appendix 1. This manuscript refers to the fourth version of the protocol.

## Methods

### Study design

Pragmatic, multicentre, randomised, 3-arm, open-label, clinical-trial enrolling patients in around 50 hospitals in Brazil. The study’s main hypothesis is that the treatment with hydroxychloroquine and hydroxychloroquine + azithromycin in patients with viral pneumonia secondary to SARS-CoV 2 improves clinical outcomes. The expected last patient last visit is end of May/early June 2020. The trial is registered with ClinicalTrials.gov (NCT04322123). The study diagram is shown in Figure 1. SPIRIT checklist is provided in the ESM appendix 2

**Figure 1.**
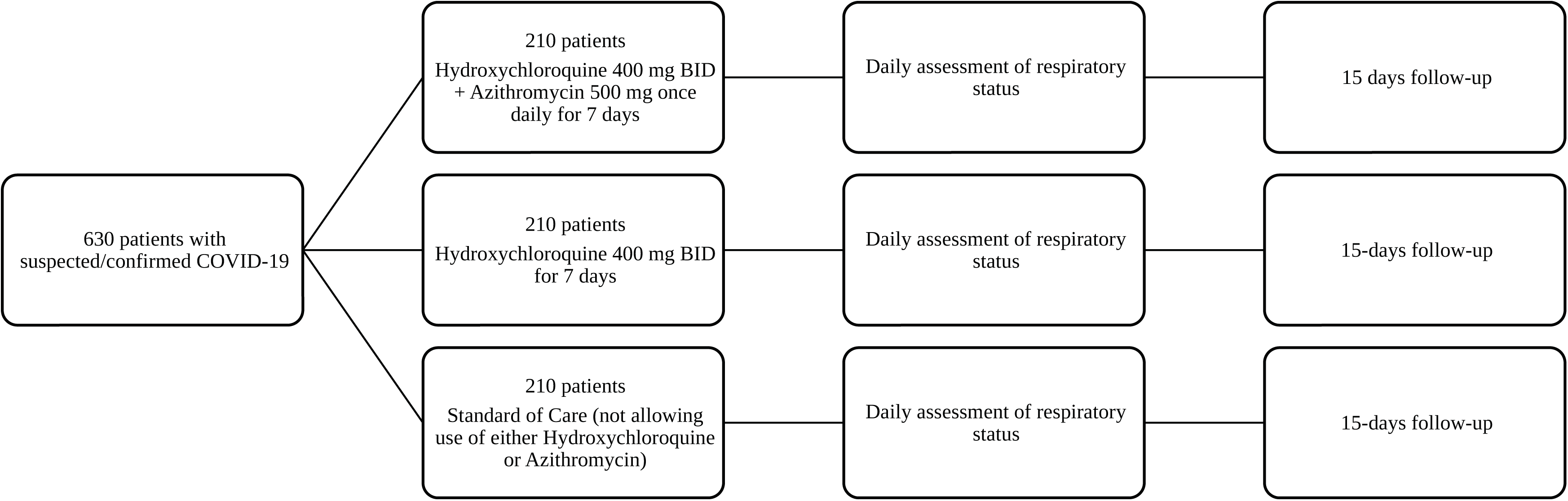
Study flowchart.

### Primary Objective

To assess the effect of the treatment with hydroxychloroquine compared to not treating with hydroxychloroquine, and the effect of combined treatment with hydroxychloroquine and azithromycin compared to treating with hydroxychloroquine only, in adult patients with confirmed COVID-19 in the ordinal outcome at 15 days:

1. not hospitalised, without limitations on activities;
2. not hospitalised, with limitations on activities;
3. hospitalised, not using supplementary oxygen;
4. hospitalised, using supplementary oxygen;
5. hospitalised, using high-flow nasal cannula or non-invasive ventilation;
6. hospitalised, on mechanical ventilation;
7. death

### Secondary Objectives

To assess the effect of treatment with hydroxychloroquine compared to not treating with hydroxychloroquine, and the effect of treating with the combination hydroxychloroquine and azithromycin compared to treating with hydroxychloroquine only, in adult patients with suspected or confirmed COVID-19 on the following secondary outcomes:

1. Ordinal outcome at day 7;
2. Need for intubation and mechanical ventilation within 15 days;
3. Need for non-invasive ventilation or high-flow nasal cannula within 15 days;
4. Length of hospital stay;
5. Hospital mortality;
6. Occurrence of thromboembolic complications (stroke, myocardial infarction, deep vein thrombosis);
7. Occurrence of acute kidney injury, defined as an increase in creatinine above 1.5 times the baseline value.
8. Number of days alive and free of respiratory support up to 15 days (DAFOR15), defined as the sum of days patients did not require supplementary oxygen, non-invasive ventilation, high-flow nasal catheter neither mechanical ventilation at 15-days. Patients that perished during the 15-day window will receive zero DAFOR15.

### Inclusion and exclusion criteria

Patients admitted to the hospital with suspected or confirmed COVID-19 will be included in the study. Inclusion and exclusion criteria are outlined in Table 1.

**Table 1.**
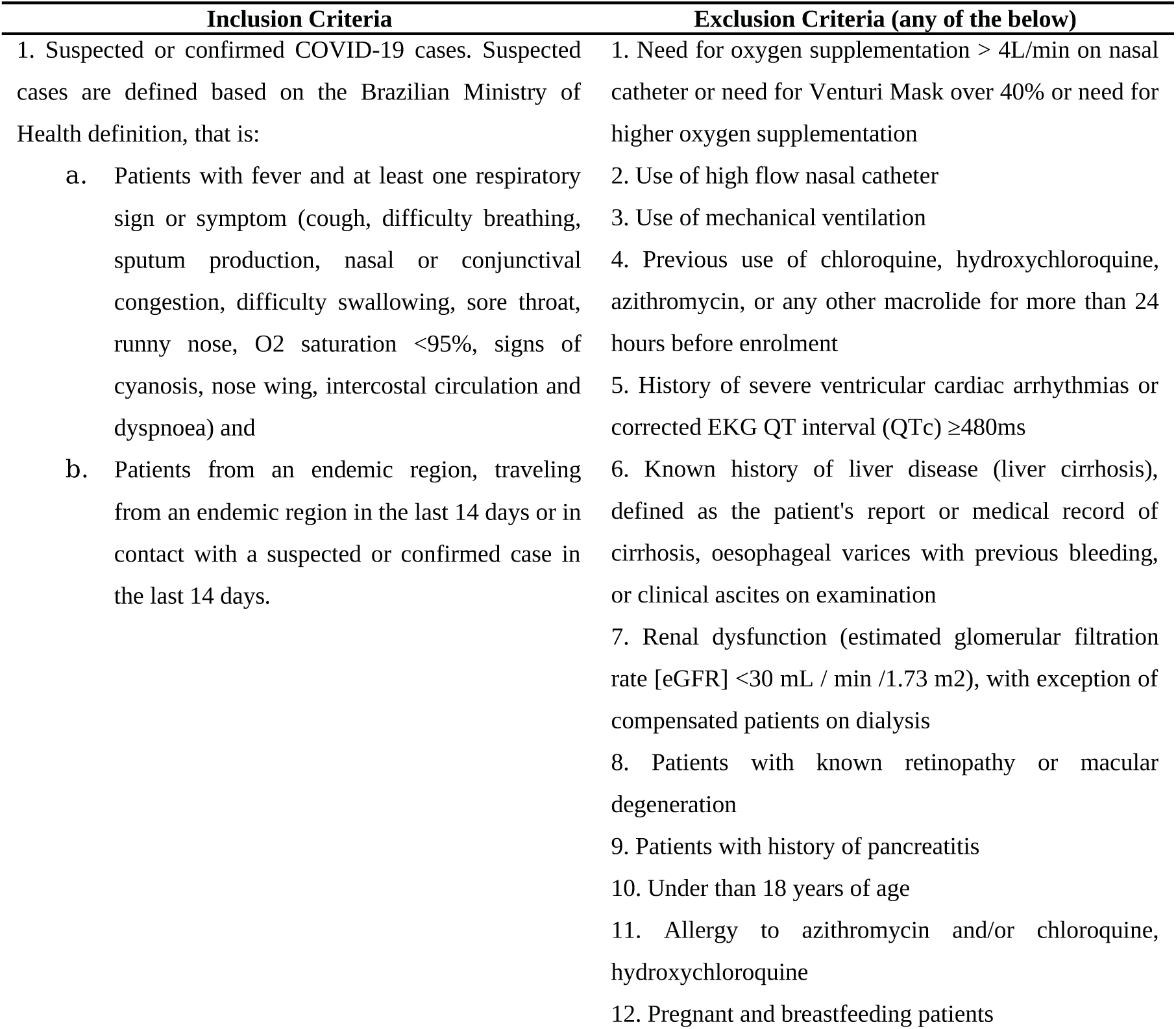
Inclusion and Exclusion Criteria.

| **Inclusion Criteria** | **Exclusion Criteria (any of the below)** |
| --- | --- |
| 1. Suspected or confirmed COVID-19 cases. Suspected cases are defined based on the Brazilian Ministry of Health definition, that is: | 1. Need for oxygen supplementation > 4L/min on nasal catheter or need for Venturi Mask over 40% or need for higher oxygen supplementation |
| a. Patients with fever and at least one respiratory sign or symptom (cough, difficulty breathing, sputum production, nasal or conjunctival congestion, difficulty swallowing, sore throat, runny nose, O<sub>2</sub> saturation <95%, signs of cyanosis, nose wing, intercostal circulation and dyspnoea) and | 2. Use of high flow nasal catheter |
| b. Patients from an endemic region, traveling from an endemic region in the last 14 days or in contact with a suspected or confirmed case in the last 14 days. | 3. Use of mechanical ventilation |
|  | 4. Previous use of chloroquine, hydroxychloroquine, azithromycin, or any other macrolide for more than 24 hours before enrolment |
|  | 5. History of severe ventricular cardiac arrhythmias or corrected EKG QT interval (QTc) ≥480ms |
|  | 6. Known history of liver disease (liver cirrhosis), defined as the patient's report or medical record of cirrhosis, oesophageal varices with previous bleeding, or clinical ascites on examination |
|  | 7. Renal dysfunction (estimated glomerular filtration rate [eGFR] <30 mL / min /1.73 m<sup>2</sup>), with exception of compensated patients on dialysis |
|  | 8. Patients with known retinopathy or macular degeneration |
|  | 9. Patients with history of pancreatitis |
|  | 10. Under than 18 years of age |
|  | 11. Allergy to azithromycin and/or chloroquine, hydroxychloroquine |
|  | 12. Pregnant and breastfeeding patients |

## Study protocol

### Randomisation Method and Allocation Concealment

The randomisation list will be generated electronically using appropriate software. Randomisation will be performed in blocks (blocks of 4 patients, 1:1:1) and will be stratified by the patient’s condition (severity group see above, that is, whether supplemental oxygen is used at this time) at the time of randomisation. Concealment of the randomisation list will be maintained through a centralized, automated, internet-based randomisation system (RedCap), available 24 hours a day. Patients who do not meet the eligibility criteria for participation in the study should not be recruited and randomised and this will be considered a major violation of the protocol.

### Blinding

Patients, investigators, and caregivers will not be blinded to the nature of the study drugs. Outcome assessors and statisticians will be blinded with respect to allocated treatments.

### Trial Interventions

Patients are randomly assigned (1:1:1) to one of three arms: control (CONT), hydroxychloroquine (HCQ) and hydroxychloroquine + azithromycin (HCQA).

The control group receive the current standard of care treatment for COVID-19, which includes daily monitoring with clinical assessment of the attending physician, routine laboratory tests (blood count, urea, creatinine, liver enzymes and bilirubin, c-reactive protein) at the discretion of the attending physician, respiratory and motor physiotherapy, surveillance of vital parameters according to the patient’s location (inpatient unit and ICU), at least once per period, which may be more frequent in some situations and institutions, addition of ventilatory support measures, such as increased oxygen flow, use of non-invasive positive pressure ventilation or high-flow nasal cannula, as recommended by the attending physician, prophylaxis of stress ulcers (if indicated) and venous thromboembolism according to the protocol of each institution and addition of other therapies such as antibiotics, corticosteroids, other antivirals (for example, oseltamivir for suspected influenza coinfection), as indicated by the attending physician. The control group does not receive hydroxychloroquine, chloroquine, azithromycin, or other macrolides.

In addition to standard care, patients in the treatment groups receive hydroxychloroquine 400 mg BID or hydroxychloroquine 400 mg BID plus azithromycin 500 mg SID, orally or enteral, for 7 days versus the standard of care (3 arms study). If patients are discharged prior to 7 days, drugs are supplied to be continued at home. If swallowing is impossible for any reason, the protocol provides for the use of the study medications via a nasogastric or nasoenteral tube. There will be no blinding at site-level, so patients and staff will know which group the patient was allocated to.

### Procedures for COVID-19 diagnosis

We require that all enrolled patients have at least one swab for COVID-19 detection through polymerase chain reaction (PCR) collected before enrolment. The clinician may enrol patients with high suspicion of COVID-19 if they fulfil other inclusion and exclusion criteria. We decided to accept cases of suspected COVID-19, instead of PCR confirmed cases only, because the results of PCR for SARS-CoV-2 are available only between 1 and 4 days in most participant sites. In this scenario, the clinician may upload other information supporting COVID-19 diagnosis to the electronic case report form (eCRF), including radiographic imaging, blood analysis and, if available, serologic tests for COVID-19. If the patient tests negative for COVID-19, he/she will be kept in the study if the physician still believes COVID-19 is the main hypothesis, otherwise treatment will be stopped but patient data will be collected until discharge and up to 15 days. For example, if the PCR is negative, but the centre maintains the suspicion due to clinical, imaging, and epidemiological data, patients continue receiving the medication until the suspicion is ruled out. Only cases with at least one RT-PCR positive for SARS-CoV-2 are classified as confirmed COVID-19. An independent committee will classify all cases with PCR tests negative for COVID into the following categories:

- Probable COVID-19: if only one PCR test for SARS-CoV-2 was performed and it is negative, and both the clinical information and chest computerized tomography (CT) are typical of COVID-19
- Possible COVID-19: if two PCR for SARS-CoV-2 were performed and are negative, and both the clinical and chest CT are typical of COVID-19; or, if only one PCR for SARS-CoV-2 was performed and is negative, and either clinical or chest CT is typical, but the other is atypical for COVID-19
- Probably not COVID-19: if two or more PCR for SARS-CoV-2 are negative, and both clinical and chest CT are not typical; or when two or more PCR for SARS-CoV-2 are negative and an alternative diagnosis explains clinical and CT findings

The main analysis study population will comprise all patients who have been randomised and have confirmed COVID (modified intention-to-treat population), but additional sensitivity analyses will be performed, considering all randomised patients and other categories (see Statistical Analysis section).

### Dose adjustment in case of adverse events

Safety adverse events are collected during trial enrolment and specific adjustments in treatment will be performed. These are resumed below, but all sites are encouraged to contact the coordinating site for instructions:

1. Nausea and vomiting: Reduce the dose of hydroxychloroquine to 400 mg daily. On recurrence, suspend temporarily. Restart in 24-48h if possible
2. Prolongation of the QT interval, defined as a corrected QT (QTc) for heart rate equal or above 480 ms: Suspend the next dose and reduce the dose to 400 mg of hydroxychloroquine per day. If QTc persists prolonged, suspend hydroxychloroquine and (if in use) azithromycin.
3. Occurrence of liver toxicity, defined as an increase in liver enzymes threefold the local normality level or any increase in serum bilirubin: Reduce the dose of hydroxychloroquine to 400 mg daily. If liver enzymes continue to rise after 24 hours of using hydroxychloroquine 1xd, discontinue the use of hydroxychloroquine.
4. Ventricular arrhythmias: stop the use of hydroxychloroquine and azithromycin.

We will also collect data on secondary safety outcomes listed below. Study treatments are to be stopped if any of those occur:

1. Severe hypoglycaemia (blood glucose ≤40mg / dL)
2. Acute cardiomyopathy (drop in ejection fraction below 40% in a patient with no previous history of ventricular dysfunction), confirmed by echocardiography
3. Hypoacusis and loss of visual acuity
4. Haematological changes such as anaemia, leukopenia, and thrombocytopenia

### Adverse event reporting and management

Adverse events are defined as any unwanted medical occurrence, including an exacerbation of a pre-existing condition, in a patient in a clinical investigation who received a pharmaceutical product. The event does not necessarily have to be causally related to this treatment. Adverse events classified as serious will be collected routinely during study visits and some special situations, even if there is no need for hospitalization, and should be reported by local investigators.

A serious adverse event is defined as any adverse event that results in death, offers immediate risk to life, results in persistent or significant disability / disability, requires or prolongs the patient’s hospitalization, results in a congenital anomaly / birth defect or must be considered serious for any other reason, if it is a major medical event that based on proper medical judgment could threaten the patient’s life or could require medical or surgical intervention to prevent one of the other results listed above.

Medical judgment should be used to determine the causal relationship of an adverse event, considering all relevant factors, including reaction pattern, temporal relationship, withdrawal or reintroduction of the drug in use. Only unexpected and not previously described serious adverse events that are believed to have a reasonable level of certainty of being related to the study medication need to be reported immediately (i.e. within 24 hours of event awareness) to the Research Institute of the HCor, local ethics committees and regulatory agencies.

The coordinating centre provides the participating site with a list of known drugs that extend the QT interval and whose use should be avoided or carefully monitored by the participating centres. This list is included in the training and is sent by email to active centres. This list is updated constantly. Caution should be taken when using any antiarrhythmic, antipsychotic, antidepressant, and other antibiotic medications, as shown in the Table 2.

**Table 2.** Suggested drug list to be avoided during the trial.

| **Antiarrhythmic** | **Antibiotics** | **Antidepressants** | **Antipsychotics** | **Others** |
| --- | --- | --- | --- | --- |
| Amiodarone | Levofloxacin | Amitriptyline * | Haloperidol | Diphenhydramine |
| Sotalol | Ciprofloxacin | Imipramine | Droperidol | Methadone |
| Procainamide | Erythromycin | Fluoxetine | Quetiapine |  |
| Ibutilide | Gatifloxacin | Sertraline | Ziprasidone |  |
| Flecainide | Moxifloxacin | Venlafaxine | Olanzapine |  |
| Digoxin | Ketoconazole | Citalopram |  |  |
|  | Itraconazole | Escitalopram |  |  |
|  |  | Bupropion |  |  |

\* and all tricyclic antidepressants in the class

### Data Collection

Information about demographic and clinical data for all patients are collected, including results of molecular tests for SARS-CoV-2, as well as the use of other therapies such as corticosteroids, antiviral agents, and other antibiotics. Patient’s clinical condition is collected daily up to the fifteenth day after enrolment or until hospital discharge. Patients discharged home before 15 days will be contacted by phone to check their vital status

Data to be collected during study visits include:

1. Admission:
  a. Age, sex, comorbidities
  b. Result of molecular tests for influenza or SARS-CoV-2
  c. Previous use of medications, such as corticosteroids, angiotensin II receptor blockers or conversion enzyme inhibitors
  d. Duration of symptoms
2. At admission and daily:
  a. Concomitant use of antibiotics
  b. Saturation, respiratory rate and inspired oxygen fraction 1x daily
    i. For patients on nasal oxygen catheter, we will collect data on oxygen flow and consider an increase of 3% over 21% for each litter per minute on nasal catheter to estimate the inspired oxygen fraction
    ii. For patients on high-flow nasal catheter: Flow and inspired fraction of oxygen
    iii. For patients on mechanical ventilation: Final positive expiratory pressure, tidal volume, inspired oxygen fraction
  c. Blood pressure and heart rate at the time of the daily assessment
  d. Need for vasopressors
  e. Laboratorial values, including liver enzymes, complete blood count (including white blood cells and lymphocytes count), creatinine, D-dimer, ferritin among others, according to the recruiting site’s routine up to day 7.
3. At telephone follow-up in 15 days: Patient status (alive/death) and whether any residual symptoms persist.

The schedule for enrolment, allocation and treatment is shown in Figure 2.

**Figure 2.**
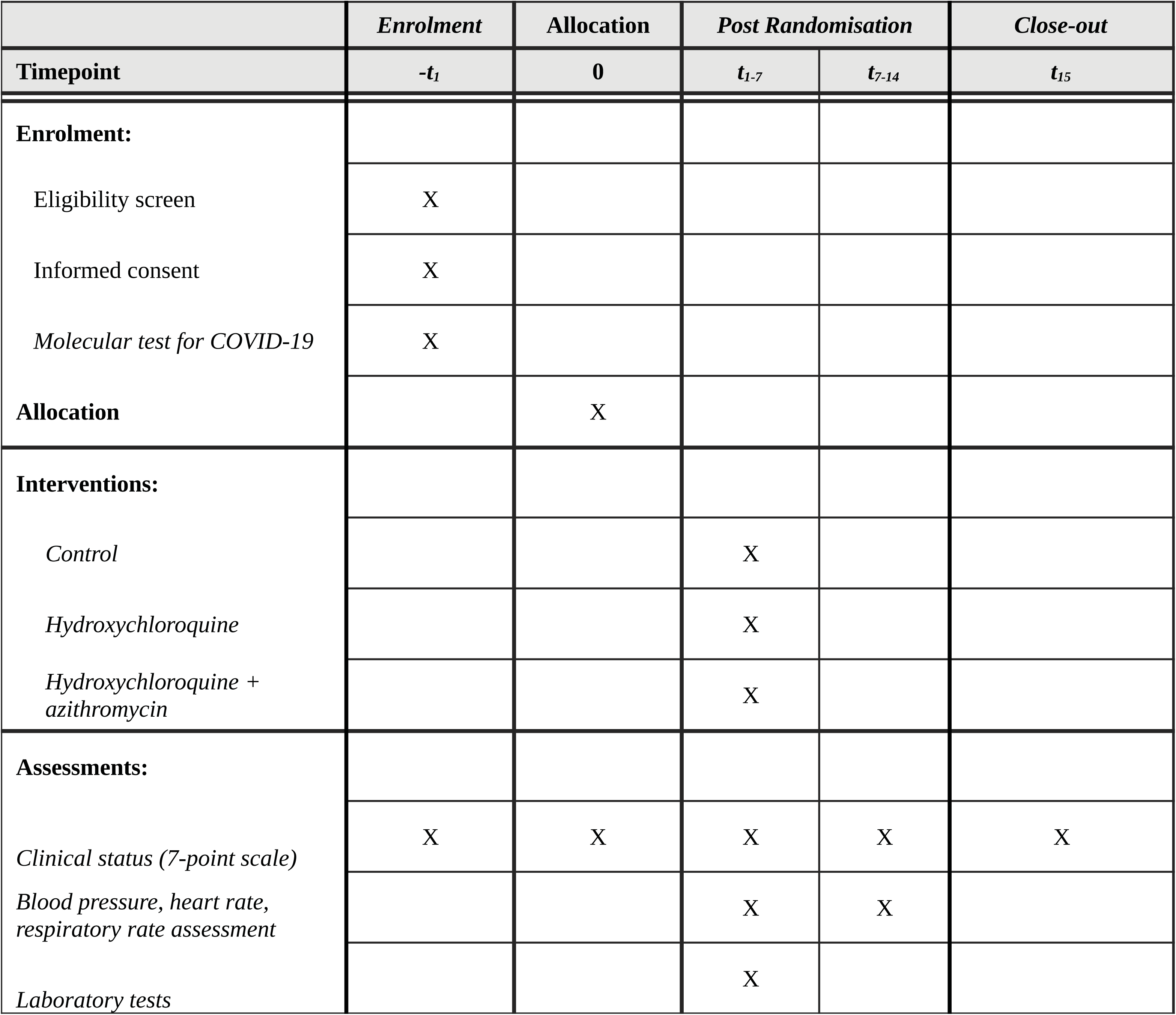
Schedule of enrolment, interventions, and assessments.

### Handling of protocol deviations

Adherence to treatment and eventual crossing of study group and protocol violations will be assessed daily. Changes in dosage as described in this protocol will not be considered deviations. Several procedures will guarantee the quality of the data, including an initial a training session before the start of the study to ensure consistency of the study procedures, availability of dedicated staff to answer site questions. We will also use statistical techniques for identifying inconsistencies and for identifying fraud and replicated records.

## Statistical Analysis

### Sample size

We will include 630 cases. We are accepting suspected cases in the study because PCR results for SARS-CoV-2 currently may take up to 5 days to be available on many Brazilian websites. Among the 630 patients, we expect to include 510 confirmed cases (with positive CRP for SARS-Cov-2), which will be the cases considered in the main analyses.

Considering an ordinal outcome with six stages with the probabilities 35%, 15%, 20%, 10%, 10%, 10%, with the expected values derived from the study of Cao [12], respectively for stages 1 to 6, under the model of proportional odds ratios for the accumulated probabilities for the outcome levels, a sample of 210 cases per arm (630 cases) has 80% power to detect a reason odds of an average of 1.76 between the arms (two by two), with a 5% significance level, with Bonferroni adjustment for multiple comparisons (alpha = 5% / 3 for each two by two comparison).

Considering the first 120 patients included in the study, we found that the distribution of the ordinal outcome in 7 levels was: 1) Out-patient, without limitation in activities, 60%; 2) Out-patient, with limited activity, 19%; 3) Patient in the hospital, without supplemental oxygen, 7%; 4) Patient in the hospital, with supplemental oxygen, 1%; 5) Patient in the hospital on non-invasive ventilation or high flow cannula, 1%; 6) Patient on mechanical ventilation, 5%; 7) Death: 7%. Assuming that this distribution represents the true distribution in the study population, with 510 patients confirmed in the main analysis, we will have 80% power to detect an odds ratio of 2.0 between the arms (two by two comparisons), with a significance level of 5 %, with Bonferroni adjustment for multiple comparisons (alpha = 5% / 3 for each comparison two by two).

### Interim Analyses

Three interim analyses are planned, the first with 120 randomised patients with the completed 15-day outcome, the second with 315 patients and a third with 504. If patient enrolment is fast, it is possible that at the time of the second or third analyses the study will have already included the total sample; interim analyses will be aborted if this happens after discussion with the data monitoring committee (DMC - see below). The DMC will use the Haybittle-Peto [13,14] stopping boundaries, considering p <0.001, to interrupt the study for safety, and an even more restrictive p-value for interruption for efficacy (p <0.0001), so we will not adjust the final values of the hypothesis test for sequential analyses. The Haybittle-Peto will be used as a reference rather than as a rigid rule. In addition, the DMC will also consider the effect on other secondary outcomes, in special adverse events, and external new evidence available during trial conduction. The p-values for multiple comparisons will be corrected by the Bonferroni method [15] in the final analysis.

### Statistical Methods

The main analysis study population will comprise all patients who have been randomised and have confirmed COVID (modified intention-to-treat population), using the group allocated as variable, regardless of the medication administered.

The primary endpoint will be assessed by mixed ordinal logistic regression considering proportional odds ratios with random intercept by site, all comparisons two by two will be performed. Binary outcomes will be assessed using a mixed logistic regression model.

Continuous outcomes will be made by generalized linear regression or mixed models for repeated variables, as appropriate. All models will be adjusted for age and need of supplemental oxygen at admission. Analyses will be performed with R software (R Core Team, 2019) [16]. Details can be found in the ESM below, Appendix 3.

### Sensitivity Analyses and Subgroup Analyses

Sensitivity analyses considering all randomised patients, independent of the results of PCR for SARS-CoV-2, shall be performed. We will also perform sensitivity analysis for the primary outcome in the following groups:

1. Definitive, probable, and possible COVID-19 patients;
2. Definitive and probable COVID-19 patients.

We should also analyse treatment effects on the primary outcome considering all patients who have confirmed COVID-19 and who received at least one dose of the medication to which they were allocated, that is, excluding patients did not take any dose of the allocated medications. For safety analyses we will consider a third population, considering the medication administered, regardless of the allocated group.

Subgroups of interest are presented in the ESM.

### Database lock

Database lock will be carried out after obtaining a 15-day follow-up for all patients and all necessary actions to obtain follow-up are carried out. All interim analyses would be made available to local regulatory agencies in Brazil. Database access will be granted only to steering committee members and statisticians before the main results are published. We plan to share data with other ongoing clinical trials for individual patient’s metanalyses.

### Protocol Amendments

In previous versions of this protocol the main analysis would follow the intention to treat principle, that is, all randomised patients would be analysed in the groups they were assigned. We decided to change the main analysis to a modified intention to treat approach considering only cases with confirmed PCR for SARS-CoV-2 for a number of reasons. First, for consistency with our primary objective, of assessing the clinical efficacy of the treatments on COVID-19 confirmed cases. Second, under the hypothesis that the treatments have beneficial effects on the primary outcome, adding non-confirmed the cases to the main analysis will decrease the estimated effect size and the power to confirm the treatment effect. Third, we anticipate that the current context of waiting days to have the PCR results will be solved in a few months. Therefore, the question “does hydroxychloroquine with or without azithromycin improve clinical outcomes for COVID-19 confirmed cases” is more relevant and will have higher external applicability than the question “does hydroxychloroquine with or without azithromycin improve outcomes of suspected COVID-19 cases”. Fourth, excluded cases without positive PCR are not associated with randomly assigned treatment, therefore bias is not an issue. Fifth, we plan to provide analyses considering all randomised cases, as well as confirmed plus probable COVID-19 cases, in the manuscript’s supplement.

### Future additions

Future versions of this protocol will all follow all necessary regulatory and ethical procedures. As the time of the writing of this protocol, we plan to add sequential swabs for a subgroup of 180 patients (60 per group) to assess qualitative viral clearance every 5 days on specific sites. We also plan to increase follow-up for longer term outcomes in future versions.

## Trial Oversight

The Steering Committee is responsible for the general supervision of the study, assisting in the development of the study protocol and preparing the final manuscript. All other study committees report to the Steering Committee. The Steering Committee members are researchers trained in the design and conduct of randomised clinical trials, intensivists, pulmonologists, cardiologists, and epidemiologists with experience in conducting multicentre randomised studies. They are listed in the electronic supplementary file. Steering committee members will act to ensure proper trial enrolment through daily contact with recruiting sites, biweekly webinars where sites are free to attend and discuss possible issues.

A Data Monitoring Committee (DMC) is formed by a statistician and two intensivists independent of the study’s investigators. The DMC is responsible for providing guidance to the Steering Committee on continuing the study as planned or stopping recruitment based on evidence that the intervention of the experimental group results in increased mortality compared to control. At the beginning of its activities, the DMC will prepare a booklet specifying the details of the formation of the DMC, its operation, meetings and interruption rules. DMC will also receive reporting of adverse events as well as regulatory agencies. After considering interim analysis, adverse events and, occasionally, external evidence, the Data Monitoring Committee should assess whether there is evidence beyond reasonable doubt that one of the interventions is clearly contraindicated for all patients. All serious or interesting adverse events will be reported to the DMC. The DMC charter is also provided in the ESM appendix 4.

## Patient and Public Involvement

There was no patient or public involvement in the design, conduct, publication or dissemination of this trial. Although desirable, patient or public involvement was felt to be not doable due to the extreme time constraint involved in the planning and execution of this trial.

## Ethics and Dissemination

All patients selected to participate in the study must sign the Informed Consent Form (ICF) before any procedure relevant to the study is carried out. Each research participant or their legal representative must provide their written consent in accordance with local requirements, after the nature of the study has been fully explained. The informed consent forms must be signed before any activity related to the study is carried out. Informed consent must comply with the principles originated in Resolution 466/12 of the National Health Council and with the guidelines of Good Clinical Practices. They can be obtained by local principal investigator or a healthcare provider trained in Good Clinical Practices delegated locally by the principal investigator. Considering the scenario of the COVID-19 pandemics, with restrictions in hospital visits from family members and decreased availability of personnel protective equipment, the site staff can obtain verbal consent upfront, with written consent obtained afterwards, or ICF can be obtained by electronic ways, such as electronic signature, from a surrogate or family members of the participants. This procedure is accordance with resolution from the Brazilian National Ethics Committee (CONEP).

Patients will be withdrawn from the study if they withdraw consent to participate in the study. In this case, the patient’s participation in the study will end and the research team will organize a final study visit to ensure patient safety. Details of the participant’s withdrawal must be documented in the source document and in the eCRF. After leaving the study, drug treatments will be determined by the patient’s attending physician. Patients can withdraw their consent to participate in the study for any reason and at any time, without prejudice to the continuity of his medical care. Patients who withdraw their consent and discontinue their participation in the study after randomisation will not be replaced by other participants. Randomised patients may also refuse to participate in specific aspects of the study or refuse to continue using the proposed drugs, without withdrawing their consent to participate. In this case, patients should be asked for permission to continue to follow them until the end of the study. Local investigators should make every effort to accommodate the needs of patients in order to maintain their participation in the study. At medical discretion, the drug (s) under study will be discontinued if the researcher considers that the discontinuation of the study drug is in the best interest of the research participant or if unexpected toxicity of the drug (s) occur.

Records of participation in this study will be kept confidential and will be accessed in a restricted way only by persons linked to the study (researchers and representatives of the study sponsor), who will transfer the clinical information to specific forms (which do not have information that can identify them). it) and verify that the study is being carried out properly. Only a number generated at the beginning of the study, the initials and / or date of birth will be used to identify the participant. The confidentiality and privacy of all information will be ensured.

This study was approved by the Brazil’s National Ethic Committee (CONEP) and National Health Surveillance Agency (ANVISA). It is registered in ClinicalTrials.gov number NCT04322123. All eventual amendments to the protocol must be approved by the IRB / CONEP System before its implementation by the participating centres. This study will be submitted for publication regardless of its results after its completion. It will also be disseminated as requested by local authorities. No professional writers will be involved during manuscript elaboration. The main paper will be authored by steering committee members plus the principal investigators of the 10-top enrolling sites which can contribute intellectually to the manuscript. The remaining principal investigators will be listed as collaborators.

## Data Availability

This is a protocol paper. No data is provided.

## Acknowledgments

None

## Author’s contribution

FGZ, ABC, RM, OB, MF, FRM conceived the trial and wrote the initial proposal. All other authors contributed for intellectually relevant content. LPD performed the sample size estimation and drafted the statistical analysis plan.

